# A Privacy-Preserving Zero-Code Conversational Statistical Analysis System for Clinical Research Using Agentic AI and Local R Execution

**DOI:** 10.64898/2026.07.30.26359367

**Authors:** Shujie Yang, Vincent Lingzhi Chen, Wee Han Ng, Siying Zhang, Sikai Qiu, Jihua Zhu, Tina Yi-Jin Hsieh, Fanpu Ji, Yee Hui Yeo

**Author notes:** Corresponding authors: Fanpu Ji, MD, PhD, Department of Hepatology, The Second Affiliated Hospital of Xi’an Jiaotong University, Xi’an, Shaanxi, P. R. China., or, Yee Hui Yeo, MD, MSc, Digestive Disease Institute, Cleveland Clinic, 9500 Euclid Avenue, Cleveland, Ohio 44195, USA. Funding: National Natural Science Foundation of China (82473291), Shaanxi Province “Three Qin Scholars” Innovation Team Project (2023001), and Fundamental Research Funds for the Central Universities (xtr062023003).

## Abstract

**Background:** Clinical data analysis typically requires statistical programming skills, whereas cloud-based artificial intelligence (AI) agents risk exposing sensitive patient records. We developed and functionally validated a privacy-preserving, zero-code conversational statistical analysis framework that translates natural-language clinical research requests into executable R workflows while strictly retaining raw patient data within local computing environments.

**Methods:** Orchestrated by the n8n engine, the system integrates the DeepSeek-Reasoner model with a Pinecone vector database for retrieval-augmented generation (RAG), grounding statistical selection in curated biostatistical guidance and R templates. Core functionalities include data schema perception, interactive data cleaning, requirements refinement, and local R code execution via a controlled command-line interface. System performance was evaluated by replicating a published prognostic model study on metabolic dysfunction-associated steatotic liver disease (MASLD).

**Findings:** All core analytical workflows — including data cleaning, multivariable Cox proportional hazards modeling, model diagnostics, and publication-ready tables and figures (e.g., baseline characteristics, Schoenfeld residuals, receiver operating characteristic curves, and forest plots) — were executed solely through natural-language dialogues without manual coding. The external large language model actively clarified analytical prompts while receiving zero row-level patient data.

**Interpretation:** Decoupling remote cloud reasoning from local code execution lowers the technical threshold for clinicians conducting data-driven research while safeguarding data privacy. This architecture provides a practical, scalable, and reproducible framework for converting natural-language clinical questions into executable statistical workflows.

**Research in context:** *Evidence before this study:* We searched PubMed, Web of Science, Embase, and IEEE Xplore for peer-reviewed research articles published from database inception up to 1 February 2026, using search terms including (“large language models” OR “agentic AI” OR “conversational AI”) AND (“clinical data analysis” OR “biostatistics” OR “R execution”) AND (“privacy-preserving” OR “local computation” OR “retrieval-augmented generation”). No language restrictions were applied. Existing clinical data analysis tools present a fundamental trade-off between analytical flexibility and ease of use. While programming languages like R and Python offer high flexibility and transparency, they require substantial statistical coding expertise. Conversely, low-code or visual workflow platforms (e.g., KNIME, LinkR) reduce coding demands but are constrained by pre-implemented, rigid analytical modules. General-purpose AI coding agents (e.g., OpenAI Codex, Claude Code) enable natural-language interaction but lack domain-grounded biostatistical frameworks to ensure methodologically sound model specification and assumption evaluation. Crucially, transmitting row-level patient data to external cloud-hosted LLM endpoints poses severe data privacy, cybersecurity, and regulatory risks (e.g., HIPAA, GDPR). To date, zero-code systems that effectively decouple cloud-based LLM reasoning from local execution of raw patient data, while incorporating domain-specific biostatistical knowledge grounding, remain scarce.

*Added value of this study:* To our knowledge, this study presents a novel, human-supervised, privacy-preserving zero-code conversational statistical analysis system that architecturally separates external LLM-assisted reasoning from local patient-level data processing. Utilizing n8n as an orchestration platform, local R execution, and Pinecone-based retrieval-augmented generation (RAG) grounded in curated biostatistical guidance and R package documentation, the system translates natural-language clinical requests into executable, reproducible R workflows. Incorporating a human-in-the-loop requirement refinement mechanism ensures that investigators retain full control over judgment-dependent decisions, such as missing-data handling and variable selection. We functionally validated the system by fully reproducing a published prognostic model study for metabolic dysfunction-associated steatotic liver disease (MASLD). Without manual programming or exposing row-level patient data to external LLMs, the system generated publication-ready baseline tables, Cox proportional hazards regressions, ROC curves, and forest plots, while reducing the analytical lifecycle from days to hours.

*Implications of all the available evidence:* Our findings demonstrate that combining external agentic AI reasoning with local, knowledge-grounded execution provides a safe, transparent, and cost-effective solution for democratizing clinical data analysis. This architecture offers a scalable and privacy-compliant blueprint for healthcare institutions seeking to empower clinicians with advanced data analytics while strictly adhering to patient data protection regulations. Future research should prioritize implementing closed-loop automated error correction, semi-automated knowledge base curation, and formal multi-center usability and statistical validity evaluations with clinical end-users.

## Introduction

In contemporary medical research, data-driven decision-making and discovery have become central forces driving progress in the field. The widespread adoption of electronic health records (EHRs) has generated large volumes of structured clinical data that can support observational research, prognostic modelling, and healthcare quality improvement.^1,2^ However, the abundance of data resources has not been equivalently translated into efficient research output. Despite increasing data availability, many healthcare institutions continue to face challenges in converting routinely collected clinical data into reproducible scientific evidence. This gap is particularly evident in institutional and multicenter EHR databases, where data acquisition often outpaces analytical capacity. Although the development of EHRs has enabled the large-scale digital collection of numerous clinical indicators and follow-up records, frontline clinicians and researchers continue to face substantial difficulties in transforming these data into evidence-based medical insights.^3,4^

One major contributor to this gap is the disconnect between clinical expertise and the technical skills required to perform modern statistical analyses. Many clinician-investigators, who are experts in understanding the clinical implications of data, lack formal training in statistical programming languages such as R or Python. Consequently, translating clinical research questions into reproducible analytical workflows often requires substantial programming support. This skill barrier not only reduces research efficiency but also limits the depth of data-driven insights that can be extracted from available datasets.^5^

To bridge this gap, both academia and industry have explored various approaches. Traditional manual programming, while powerful, demands substantial time investment for syntax learning and debugging, resulting in a steep learning curve. Graphical user interface (GUI)-based statistical software such as SPSS, SAS Enterprise Guide, and GraphPad Prism, reduces operational difficulty through menu-driven interfaces but often provides limited flexibility for highly customized analytical workflows, which may restrict complex or iterative analyses. Low-code or no-code platforms attempt to empower non-technical users by encapsulating underlying code logic; however, their analytical capabilities are typically constrained by predefined functional modules and are frequently constrained by predefined analytical modules and may not readily support investigator-specific analytical workflows.^7–8^

The emergence of large language models (LLMs) for code generation has opened a novel pathway, yet current implementations exhibit inherent limitations in medical statistical tasks: (1) Limited awareness of local datasets: Stand-alone LLM interfaces do not inherently maintain reliable awareness of local dataset schemas, variable definitions, or analytical state. The code they generate often contains “hallucinations” including fabrication of column names, inappropriate package selection, and generation of non-executable code. (2) Inability to execute and verify automatically: Users must manually copy, paste, execute, and debug generated code, breaking workflow continuity. (3) Contextual discontinuity: In multi-step complex analyses, LLMs struggle to maintain consistent context across multiple conversation turns. (4) Privacy and governance concerns: Many high-performance LLM services rely on cloud-based inference, raising governance concerns when patient-level information is transmitted outside institutional environments. This conflicts with the stringent data privacy protection requirements in medical research.^9–10^ (5) Limited reproducibility of generated analytical workflows: Variability in prompt interpretation and code generation may produce inconsistent analytical workflows across repeated sessions.^11^ Recently, several cutting-edge architectures have attempted to address these challenges. The Model Context Protocol (MCP), introduced by Anthropic, establishes a standardized open interface for connecting LLMs with local databases;^12^ however, MCP primarily facilitates communication between models and external tools and does not itself ensure methodological validity, statistical appropriateness, or analytical reproducibility. Similarly, modular instruction architectures such as Skill.md can improve performance on specific tasks but lack systematic design for the complex, long-chain workflow from data cleaning to hypothesis testing.^13^

Given these limitations, a privacy-preserving artificial intelligence (AI)-assisted analytical workflow that can generate executable statistical code for clinicians who lack dedicated programming or statistical expertise, while ensuring patient data privacy through fully localized operation, could substantially accelerate clinical research productivity, improve analytical reproducibility, and facilitate broader participation in data-driven healthcare innovation. Such a system should preserve human oversight of study design, variable selection, missing-data management, and model interpretation rather than fully automating analytical decision-making. This potential is especially meaningful in resource-limited environments where access to dedicated statistical expertise is constrained. Critically, such a system should be positioned as an intelligent collaborator that augments rather than replaces professional statisticians.

The emergence of agentic AI offers a promising architectural paradigm to realize this vision, yet its reliability and applicability in highly specialized and rigorous medical research scenarios require further empirical validation.^14^

In this study, we aimed to develop and validate a human-supervised conversational statistical analysis system that combines agentic workflow orchestration, retrieval-augmented generation, and local R execution. The resulting analytical system will empower clinicians to independently perform routine analytical tasks.

## Methods

### Environment Configuration and Deployment

The analysis environment was deployed on a local workstation using Node.js and n8n (v2.14.2) as the workflow orchestration engine and R (v4.7.3) as the statistical computing backend. wenty-three preinstalled R packages supporting data manipulation, statistical modelling, survival analysis, machine learning, and visualization (e.g., tidyverse, survival, glmnet, pROC, ggplot2) was used. n8n orchestrates workflow execution and data exchange, whereas R scripts are executed locally through the Rscript interface to perform statistical analyses and generate graphical outputs. Communication between n8n and R occurs through predefined local file paths and command-line invocation, ensuring that all patient-level data processing and statistical computation remain local (Figure 1).

**Figure 1.**
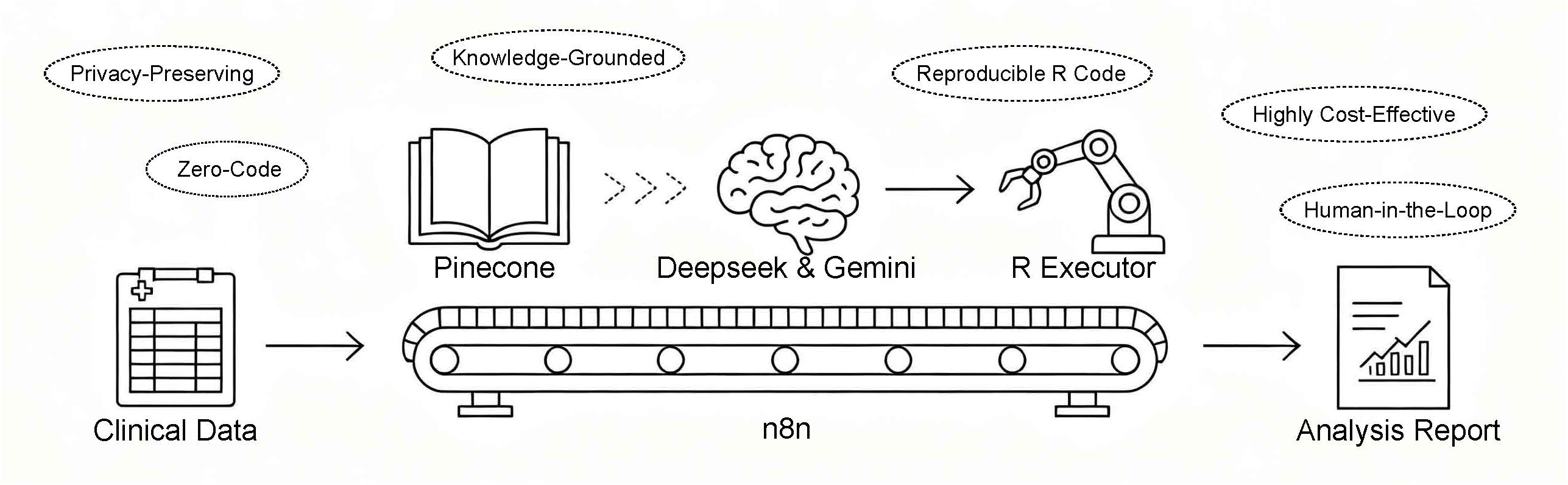
Conceptual framework and core highlights of the conversational clinical statistical analysis system. The schematic illustration presents an end-to-end overview of the system architecture conceptualized as an automated analytical pipeline. The process initiates with the ingestion of raw Clinical Data and culminates in the generation of a comprehensive, publication-ready Analysis Report. The entire pipeline is centrally orchestrated by the n8n workflow engine, represented as the underlying conveyor belt that manages the sequential data and task flows. To eliminate the technical barriers for clinicians, the system provides a Zero-Code conversational interface. The core intelligence relies on large language models (Deepseek & Gemini) acting as the reasoning engine. To mitigate model hallucinations and guarantee that the system is Knowledge-Grounded, a Pinecone vector database provides retrieval-augmented generation (RAG) by injecting biostatistical guidelines and package documentation into the LLM prompt. The refined analytical intent is then translated into code and executed via a local R Executor (represented as the robotic arm), which guarantees the production of transparent and Reproducible R Code. Crucially, the architecture is Privacy-Preserving, as raw patient-level records are confined to the local environment, and incorporates a Human-in-the-Loop supervisory design where critical clinical judgments require investigator validation. By utilizing open-source tools and API-based inference, the system offers a Highly Cost-Effective alternative for data-driven medical research. Abbreviations: LLM, Large Language Model; RAG, Retrieval-Augmented Generation.

### Data Privacy and Local Processing

The system was designed to prevent transmission of row-level patient records to external LLM services. Metadata transmitted to the LLM included variable names, inferred data types, missingness summaries, and high-level distribution characteristics. Complete patient-level records remained within the local workstation throughout the analytical workflow. The LLM is accessed via API only with this anonymized structural information, and all generated R code was executed locally within the R environment. This architecture minimizes external exposure of patient-level clinical data while allowing LLM-assisted code generation and workflow orchestration.

### System Architecture Overview

To bridge the gap between natural-language clinical queries and the execution of reproducible statistical workflows without exposing sensitive patient records to external servers, the proposed system implements a four-tier Systematic Layered Architecture (Figure 2). This architecture segregates the user experience, logic orchestration, AI reasoning, and data computation into independent operational domains:

**Figure 2.**
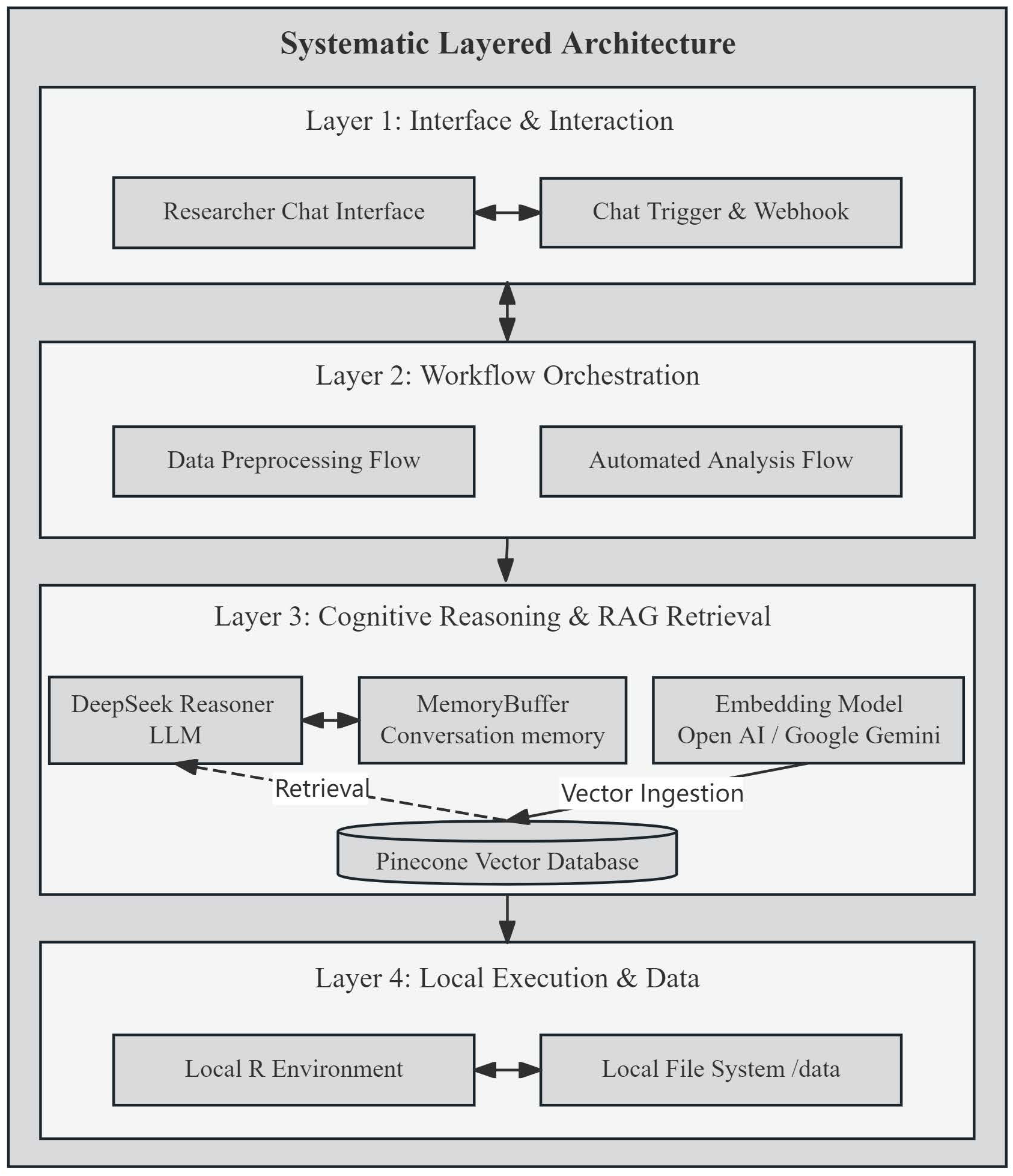
Detailed schematic and data-flow pathways of the systematic layered architecture. The architectural diagram illustrates the four-tier modular layout of the platform, highlighting the segregation of cloud-assisted reasoning from local sandboxed computation. The vertical flow is initiated at Layer 1 (Interface & Interaction), where the Researcher Chat Interface communicates bidirectionally (indicated by the horizontal double-sided arrow) with the Chat Trigger & Webhook to capture user intent. This event is passed down via a vertical solid arrow to Layer 2 (Workflow Orchestration), which visually separates the pipeline into two parallel functional containers: the Data Preprocessing Flow and the Automated Analysis Flow. A vertical solid arrow points further down to Layer 3 (Cognitive Reasoning & RAG Retrieval), detailing the knowledge-grounded generation loop. Here, the horizontal double-sided arrow between the Deepseek Reasoner LLM and MemoryBuffer denotes real-time conversational memory synchronization. The solid downward arrows show the Vector Ingestion pathway from Google Gemini embedding algorithms into the centralized Pinecone Vector Database, while the diagonal dashed arrow marked “Retrieval” explicitly represents the context-dependent data stream that restricts the LLM’s reasoning within biostatistical boundaries. Finally, the downward solid arrow leads to Layer 4 (Local Execution & Data), defining the data privacy boundary. The horizontal double-sided arrow at the bottom signifies direct, isolated local read/write pathways between the Local R Environment and the Local File System (/data), illustrating that row-level data streams remain completely encapsulated within the local workstation. Abbreviations: LLM, Large Language Model; RAG, Retrieval-Augmented Generation.

#### Layer 1: Interface & Interaction

This outermost layer establishes a conversational gateway for clinicians. It captured natural-language analytical requirements and converted them into system-readable events, enabling a zero-code interactive experience without requiring technical knowledge of underlying APIs.

#### Layer 2: Workflow Orchestration

Serving as the central controller of the system, this layer managed sequential and conditional execution logic using localized workflow orchestration. The orchestration logic is divided into two major operational pipelines: a data preprocessing track tasked with dataset structural perception and anomaly cleansing, and an automated statistical analysis track dedicated to analytical requirement diagnosis and output generation.

#### Layer 3: Cognitive Reasoning & RAG Retrieval

This layer governs requirement refinement, biostatistical decision-making, and script synthesis. It pairs a reasoning-focused LLM with conversation memory to maintain clinical context across multi-step dialogues. To systematically mitigate model hallucinations, a RAG framework is embedded within this layer, constraining the LLM’s code generation within verified biostatistical knowledge bases and standardized R coding specifications.

#### Layer 4: Local Execution & Data

Acting as the ultimate security perimeter for patient privacy, this foundational layer enforces absolute local data isolation. The synthesized statistical scripts are dispatched to a sandboxed local environment where all patient-level data transformations, mathematical modeling, and figure generation are executed entirely offline against the local storage, guaranteeing that raw row-level patient data never leave the investigator’s local infrastructure.

### Core Analysis Pipeline

The analytical process is implemented as two interacting workflows (Figure 3).

**Figure 3.**
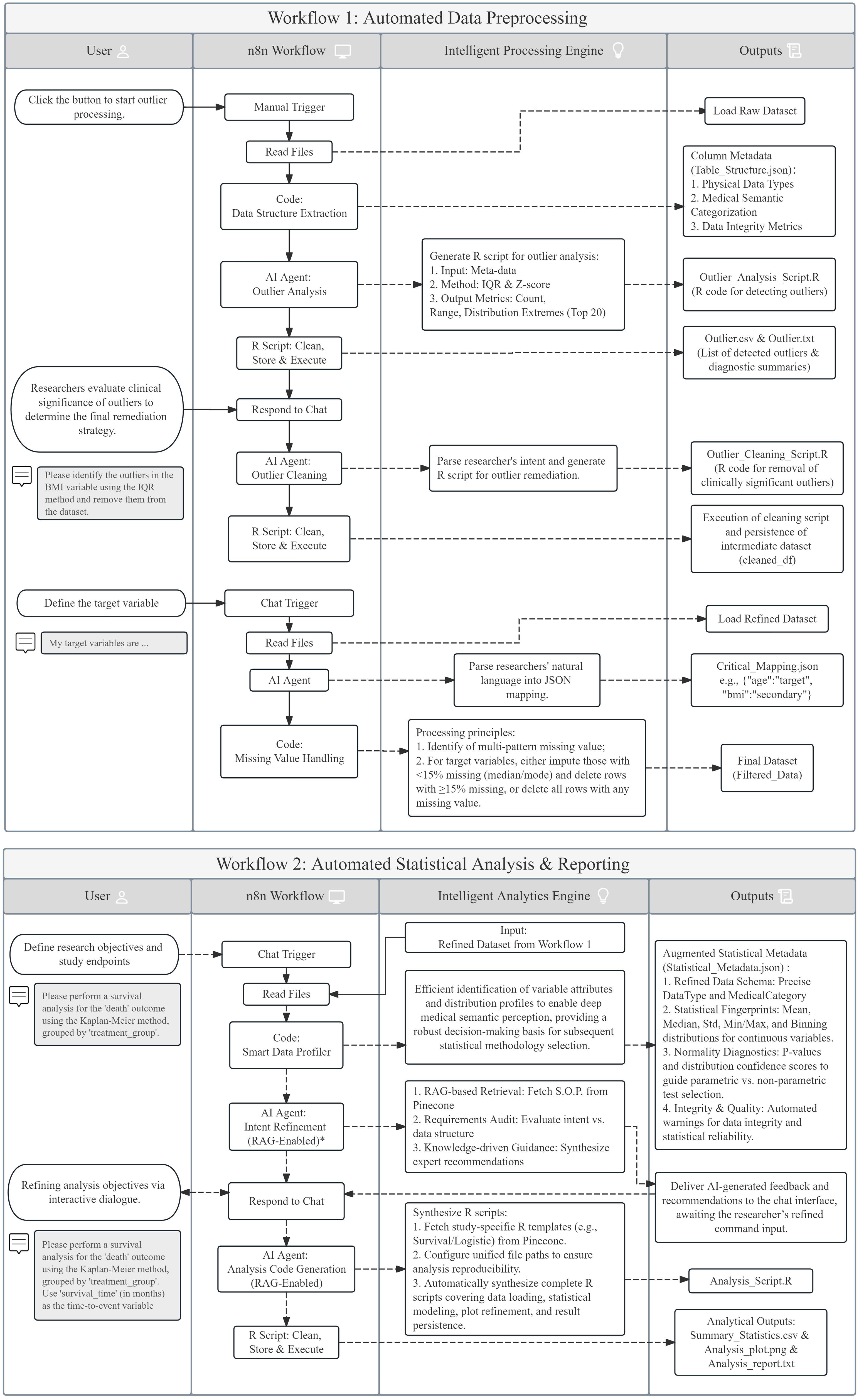
Operational architecture and logical data-flow schema. Workflow 1: Agentic AI–Driven Data Preprocessing and Outlier Handling with Human– AI Interaction Loops; Workflow 2: Retrieval-Augmented Generation (RAG) Pipeline for Analytical Requirement Refinement and Automated Statistical analysis.Abbreviations: AI, Artificial Intelligence; BMI, Body Mass Index; IQR, Interquartile Range; JSON, JavaScript Object Notation; LLM, Large Language Model; Max, Maximum; Min, Minimum; RAG, Retrieval-Augmented Generation; S.O.P., Standard Operating Procedure; Std, Standard Deviation.

Human–AI Interaction Loop Workflow 1 – Local Data Profiling and Preprocessing: After a researcher uploads a CSV file, a Code node extracts the table structure and identifies potentially extreme observations using both interquartile range (IQR)-based and Z-score-based screening methods. A summary of resulting statistics is generated. The researcher may then issue natural language commands (e.g., “winsorize extreme creatinine values”) to trigger an AI Agent that generates and executes the corresponding R code.

Human–AI Interaction Loop Workflow 2 – Automated Analysis: Triggered by a natural-language analytical query, this workflow first generates a detailed data dictionary via automated schema and distribution profiling. An AI Agent refines the analysis requirement by consulting the Pinecone knowledge base and conducting a multi-turn dialogue with the researcher. After investigator confirmation of the analytical plan, a second AI Agent produces executable R code, which is cleaned, written to disk, and executed locally via Rscript. All statistical tables and figures are saved to a designated output directory.

### Implementation of the Core Algorithm

The system automates the traditional epidemiological analysis workflow (data perception, data cleaning, requirement refinement, and code execution) through four integrated components.

#### Intelligent Data Perception and Interactive Data Cleaning

Researchers can launch this workflow by specifying the variables to be included in the analysis (i.e., target variables) via natural language. The system then automatically loads the CSV file and scans the dataset. Relying on predefined clinical terminology dictionaries and heuristic classification rules, it automatically infers the storage type of each variable (continuous, binary, categorical, ordinal, date/time, or identifier) and determines its applicable analytical role (continuous, categorical, ordinal, or identifier). Meanwhile, the system identifies all missing values based on the missing value pattern library and processes them following standard schemes. Our system provides two predefined missing-data management options for investigator selection: 1. Perform simple imputation for variables with less than 15% missing data (median imputation for continuous variables, mode imputation for categorical variables); 2. delete all cases with missing values for variables with a missing rate of 15% or higher. The 15% threshold serves as a configurable default parameter within the proof-of-concept workflow and can be adjusted freely by researchers. Directly delete all cases that contain any missing value in any target variable.

Non-target variables (secondary variables) remain unprocessed throughout the entire procedure.

For outlier handling, after identifying variable types, the system applies both the interquartile range (IQR) method and the Z-score method to all continuous variables simultaneously to detect outliers and generates an outlier detection report. Investigators may input additional natural-language cleaning instructions (e.g., “Winsorize the outliers for creatinine” or “delete rows where total bilirubin is an outlier”). The AI Agent parses these instructions into executable R code, runs the code locally, and saves the cleaned dataset on the local machine.

#### AI-Assisted Requirement Refinement

The researcher expresses analytical intent in natural language (e.g., “compare liver function indicators between the survival group and the death group”). The AI Agent compares the analytical request against the available data dictionary and proposes candidate analytical approaches, retrieves relevant methodological guidance and diagnostic considerations from the Pinecone knowledge base. It then returns supplementary recommendations, such as suggesting non-parametric tests when the data deviate from normality. Once the researcher confirms or adjusts the suggestions, the investigator-confirmed structured analytical plan is submitted through the dialogue interface, and the system proceeds to generate the corresponding code.

#### Intelligent Code Generation and Execution

After the analytical plan is confirmed, a dedicated AI Agent generates an executable R analysis script. The system prompt strictly constrains the agent to use fixed local file paths, automatically perform variable type conversions (e.g., converting categorical variables to factors), and select appropriate statistical tests and visualization functions. The generated code is cleaned of Markdown formatting and executed via the Rscript command in the local R environment. All outputs (tables and figures) are saved to a designated local directory, while maintaining local execution of all patient-level statistical computations.

#### Construction of the Clinical Research and Biostatistics Knowledge Base

The Pinecone vector database stores five categories of domain knowledge: R programming standards, publication-style figure templates, biostatistical methodology guidance, R package recommendations, and interactive requirement templates. During both requirement refinement and code generation, the AI Agent is mandated to query this knowledge base. The query text is embedded into a vector, and the most relevant knowledge snippets are retrieved by cosine similarity and injected into the generation context. This RAG design mitigates the “hallucination” problem of large language models in medical statistical tasks at two levels. First, the verified statistical strategies and code templates in the knowledge base provide curated reference material reviewed for methodological and coding relevance. This substantially reduces the probability of fabricating nonexistent R functions or erroneously selecting statistical methods due to a lack of professional knowledge. Second, when a complex analytical task is detected (e.g., drawing a Kaplan-Meier curve or building a Cox regression model), the system proactively retrieves parameter templates from the knowledge base and guides the researcher with structured questions to supply key parameters. This step reduces the likelihood of unsupported assumptions when analytical information is incomplete. Although RAG cannot eliminate all coding or methodological errors, it constrains code generation using curated statistical resources and standardized analytical templates.

### Functional Validation Through Replication of a Published Study

Functional validation was performed by replicating the principal analytical workflow of a published prognostic model study in MASLD.^15^ Before workflow execution, dataset harmonization, including variable renaming and value standardization, was performed manually using the official manual of National Health and Nutrition Examination Survey (NHANES) database. Furthermore, population screening was performed manually based on inclusion criteria (individuals aged 20–70 years with MASLD defined by a MAF-5 score ≥ 0) and exclusion criteria (high alcohol consumption, defined as ≥140 g/week for females or ≥210 g/week for males, or missing data). Therefore, validation focused on the analytical workflow rather than automated cohort construction.

Validation endpoints included successful completion of requested analytical tasks, generation of executable R code, reproduction of the principal statistical outputs reported in the reference study, and confirmation that row-level patient records were not transmitted in outbound requests to the external LLM.

## Results

### System implementation and workflow completion

The final system comprised two interconnected workflows: (1) Automated Data Preprocessing (Figure S1a, b), and (2) Automated Statistical Analysis & Reporting (Figure S1c). Functional validation was conducted by reproducing the principal analyses of a published metabolic dysfunction-associated steatotic liver disease (MASLD) prognostic model study.^15^ The investigator initiated and confirmed analytical decisions through natural-language interaction without manually writing or executing any R code. A representative dialogue sequence is shown in Figure 4; complete prompts, responses, generated code, and output files are provided in Supplementary Appendix E.

**Figure 4.**
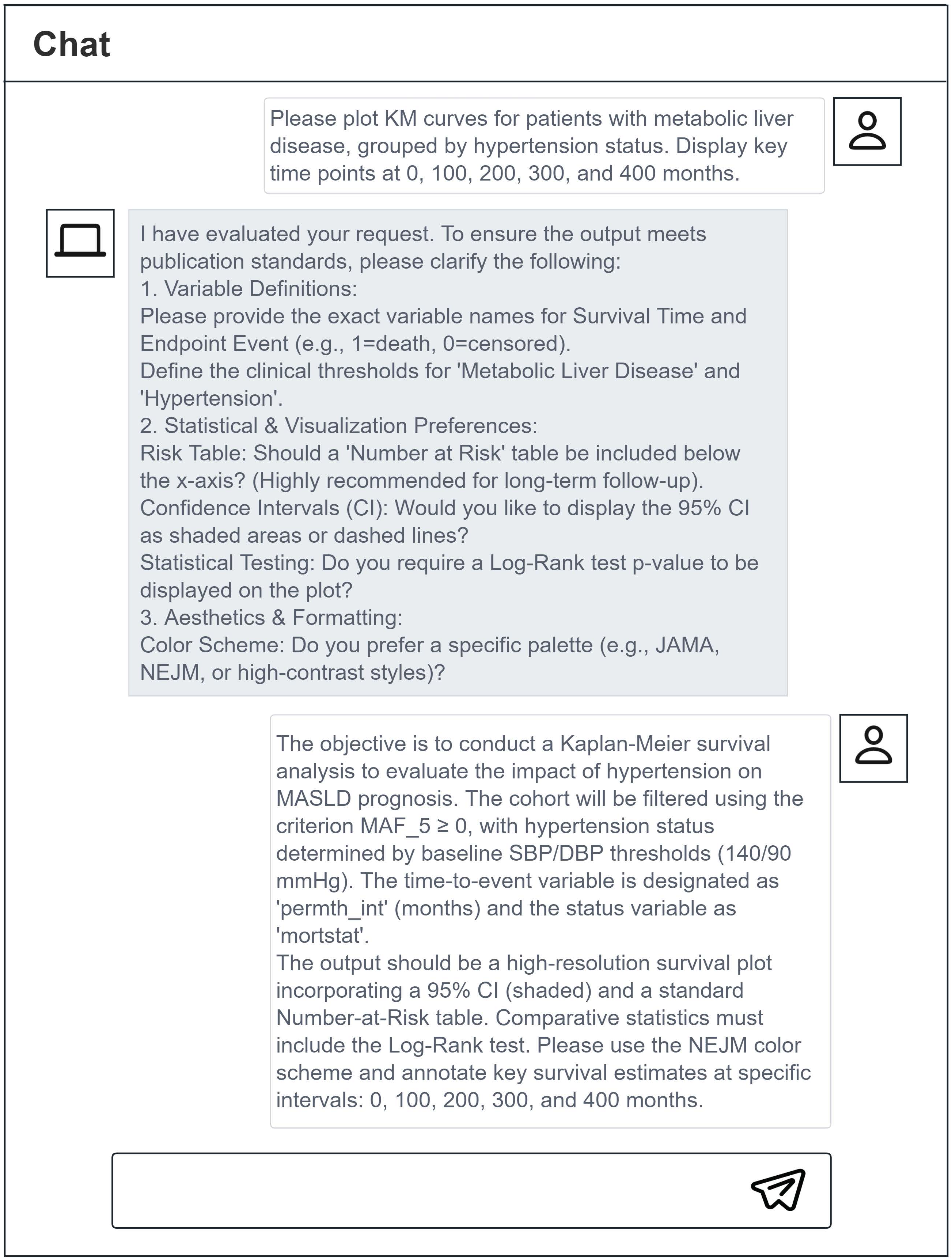
Schematic diagram of the complete dialogue in the analysis process.

### Replication of the published analysis

The system performed all prespecified analytical steps: (i) Kaplan – Meier estimation, (ii) descriptive analyses with univariable Cox regression, (iii) multivariable Cox regression and model performance evaluation, and (iv) ROC curve generation. The resulting tables, Kaplan–Meier curves, and forest plots are provided in the Supplementary Material (Tables S1–S4 and Figure S2–S3); and the ROC curve is displayed in Figure 2 of the main text. Three tasks were executed successfully on the first attempt, whereas the remaining task required an iterative workflow reuse. Specifically, the initially refined analysis commands were re-submitted to further normative optimization, thereby generating a more comprehensive and accurate analytical directive that ultimately enabled the AI agent to produce rigorous executable code.The final model, determined by stepwise regression, retained 13 variables: age, body mass index, waist circumference, systolic blood pressure, diastolic blood pressure, total cholesterol, fasting blood glucose, aspartate aminotransferase, alanine aminotransferase, gamma-glutamyl transferase, platelet count, total bilirubin, and creatinine. This set differs slightly from that of the original study, which included sex and alkaline phosphatase instead of body mass index and platelet count, which is a discrepancy that may be attributed to differences in the initial candidate variable pools entered into the stepwise selection procedure. Our final model achieved a C-index of 0.778 (95% CI: 0.762-0.794) (Table S4); the 5-, 10-, and 20-year time-dependent area under the receiver operating characteristic curves (AUCs) were 0.786, 0.776, and 0.820, respectively (Figure 5).

**Figure 5.**
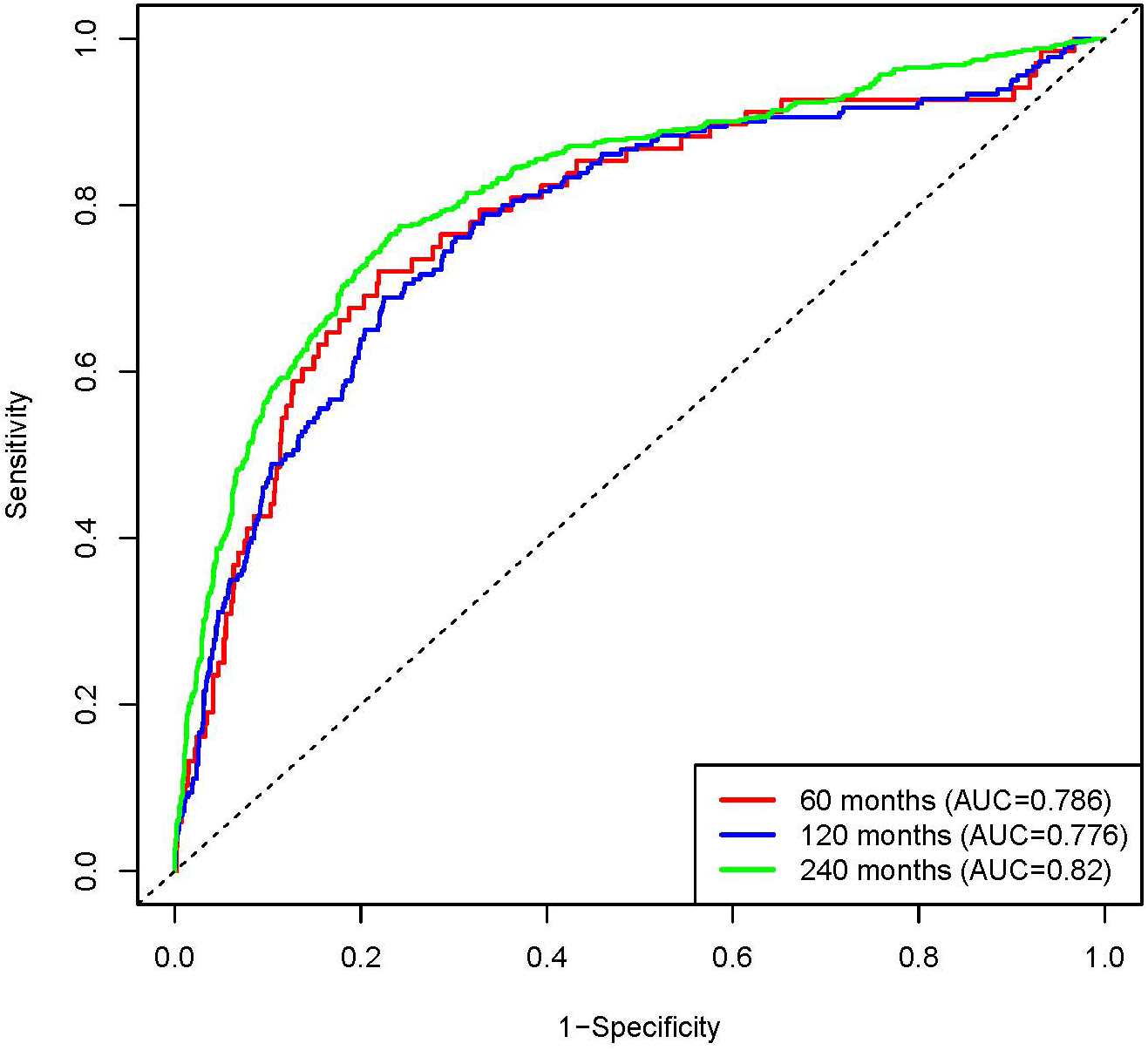
Time-dependent ROC curves for predicting all-cause mortality in MASLD patients at 60, 120 and 240 months. Time-dependent receiver operating characteristic (ROC) curves of the final multivariate Cox regression model for predicting all-cause mortality among MASLD patients at different follow- up time points. The area under the curve (AUC) values were 0.786 for 60- month survival, 0.776 for 120- month survival, and 0.820 for 240- month survival, respectively. The diagonal dashed line represents the reference line with an AUC of 0.5 (no predictive ability). Abbreviations: ROC, receiver operating characteristic; AUC, area under the curve.

### Analytical fidelity

The reproduced effect estimates were consistent with those reported in the reference study. The absolute relative differences in hazard ratios ranged from 0.003 to 0.183, and the difference in time-dependent AUCs remained below 0.034.^15^ The 5-year AUC from our system was 0.786, compared with 0.764 in the original study.

### Processing time and reproducibility

The complete workflow, from data ingestion to final output generation, required approximately 3 hours. Importantly, re-execution of the saved R scripts in the same software environment produced identical numerical outputs. No manual modification of the final R scripts was required.

### Data Privacy and Security

All patient-level data processing and statistical computation were performed within the local workstation, and no data were transmitted to external servers. To further protect participant privacy, the raw data were processed through a dedicated structure-extraction node that generated column-level summaries (e.g., data types, missing rates, distributional characteristics) without exposing any complete patient records. This ensured that the LLM, which handled only the aggregated structural metadata, could not access sensitive patient information (Figure S4).

## Discussion

In this proof-of-concept study, we developed and functionally validated a human-supervised, zero-code conversational statistical analysis system that combines agentic workflow orchestration, retrieval-augmented generation, and local R execution. Using natural-language interaction, the system completed the prespecified analytical workflow of a published study, including data-quality assessment, survival analysis, Cox regression, model diagnostics, and generation of publication-formatted tables and figures. Patient-level data processing and statistical computation remained within the local workstation, while the external LLM was restricted to analytical requirement refinement and R code generation using predefined dataset metadata and retrieved statistical guidance. The principal contribution of this system is therefore not simply automated code generation, but the architectural separation of external LLM-assisted reasoning from local patient-level computation within an investigator-supervised and auditable workflow.

### Comparison with Existing Clinical Data Analysis Approaches

Existing approaches to clinical data analysis often involve a trade-off between analytical flexibility and ease of use. Programming environments such as R and Python provide extensive flexibility and transparency but require proficiency in statistical coding. In contrast, graphical and low-code platforms reduce programming demands but generally require investigators to select appropriate methods, configure analytical parameters, and interpret statistical assumptions.

Visual workflow platforms such as KNIME and Galaxy enable users to construct transparent and reusable analytical pipelines with limited manual coding. However, investigators must still identify the appropriate analytical modules, specify parameters, evaluate assumptions, and interpret the resulting outputs.^16–19^ Healthcare-oriented low-code platforms such as LinkR further improve the accessibility of data manipulation and statistical analysis. However, their functionality remains dependent on the analytical modules and workflows implemented within the platform.^7^ General-purpose coding agents, including OpenAI Codex and Claude Code, permit natural-language interaction with source code and can generate, execute, and revise analytical scripts. However, these tools are primarily designed for general software-engineering tasks and do not inherently provide a curated framework for clinical epidemiologic study design, biostatistical method selection, or assessment of model assumptions.^20–23^ Biomedical AI platforms such as Polly Co-Scientist provide greater domain specialization but are designed to augment broader biomedical research workflows rather than serve as a transparent, general-purpose system for conversational clinical statistical analysis. In addition, workflows that transmit patient-level data to externally hosted models require appropriate contractual, regulatory, privacy, and cybersecurity safeguards. These considerations identify a need for a knowledge-grounded conversational system that reduces the requirement for manual programming while preserving investigator control, statistical transparency, and local processing of patient-level data.

### Architectural and Methodological Contributions

The proposed system addresses these gaps through the integration of conversational requirement refinement, retrieval-grounded R code generation, local statistical execution, and preservation of an auditable analytical record. First, the system allows investigators to initiate and refine multistep analytical tasks through natural-language interaction without using R code. It translates investigator-specified decisions regarding data cleaning, variable selection, statistical modelling, model diagnostics, and output formatting into executable R scripts and publication-formatted analytical outputs. Second, the requirement-refinement process presents proposed analytical specifications to the investigator for review and confirmation before code execution. This human-in-the-loop design preserves investigator control over judgment-dependent decisions, including missing-data management, outlier handling, covariate selection, model specification, and interpretation of diagnostic findings. Third, the architecture separates external LLM-assisted reasoning from local patient-level computation. Row-level data transformations, statistical modelling, and generation of final outputs are performed within the local workstation, whereas the external LLM receives only the predefined metadata and aggregate information required to refine the analytical request and generate R code. Fully local deployment of an open-weight LLM represents an alternative approach that can avoid external model inference. However, it may require substantial computing infrastructure, model deployment expertise, maintenance, and local validation. Moreover, local hosting addresses the location of data processing but does not itself prevent hallucination.

Fourth, the system combines a free self-hosted n8n Community Edition, the open-source R statistical environment and R packages, locally hosted embedding models, and usage-based external services for LLM inference and vector retrieval. Although it is not entirely open-source or cost-free, this architecture avoids the licensing costs of commercial statistical software. Furthermore, because the LLM API is used exclusively for text generation (R code and natural language dialogue), the per-analysis token cost is substantially lower than that of tools relying on multimodal models or extensive cloud computation. These combined cost advantages make the system particularly suitable for resource-limited settings, where dedicated statistical expertise and commercial analytical tools are financially inaccessible.

A central design objective was to mitigate the risk of hallucinations (unsupported, non-executable, or statistically inappropriate outputs) from LLMs. The system addresses this risk through three complementary mechanisms. First, local schema profiling provides the LLM with predefined information regarding variable names, storage types, analytical roles, missingness, and distributional characteristics, reducing reliance on assumed dataset structures. Second, retrieval-augmented generation (RAG) grounds method selection and code construction in curated biostatistical guidance, R package documentation, reporting recommendations, and validated code and visualization templates. Third, all numerical analyses are delegated to the R statistical environment, and the resulting scripts and outputs remain available for investigator inspection, execution review, and reproduction. Together, these mechanisms constrain the generation process and reduce the likelihood of fabricated variables, nonexistent functions, and unsupported analytical assumptions. Nevertheless, retrieval grounding does not independently validate the appropriateness of the selected method, the correctness of the generated code, or the interpretation of the resulting estimates.

Finally, the system accelerates the translational cycle of clinical research by compressing workflows that traditionally require days into hours. Preservation of the final R scripts, data-processing decisions, package versions, and analytical outputs supports transparency and computational reproducibility once the code has been finalized. Regarding reproducibility, it should be distinguished at the code-execution and LLM-generation levels. A frozen R script can reproduce identical numerical results when executed with the same data, software versions, and random seeds. In contrast, repeated LLM interactions may generate different analytical plans or code because of model stochasticity, model updates, prompt variation, and differences in retrieved knowledge. Reproducibility of the complete conversational workflow therefore requires preservation of the model identifier, system and user prompts, retrieved knowledge snippets, generated code, package versions, random seeds, and execution logs.

### Limitations and Future Directions

Several limitations of the current implementation warrant acknowledgment. The non-deterministic nature of LLM code generation means that syntax errors or suboptimal statistical choices may still occur. The current system captures execution errors and terminates, but lacks an automated error-correction feedback loop, which is an important direction for future development. Dependence on external LLM APIs introduces vulnerability to network latency and service interruptions. The initial construction and ongoing maintenance of the knowledge base require manual expert curation; future work will explore semi-automated literature mining approaches to reduce this burden. The system currently supports only structured tabular data and does not yet extend to unstructured data types such as medical images or free-text clinical notes. Furthermore, user input with high ambiguity or internal contradictions may exceed the capacity of the current requirement clarification mechanism; a more robust degradation and re-prompting strategy is needed. Finally, the validation presented in this paper is primarily technical; formal usability studies with clinical end-users are planned to empirically evaluate task completion efficiency, user satisfaction, and learning cost.

In conclusion, this proof-of-concept study demonstrates the feasibility of using a human-supervised, retrieval-grounded conversational system to translate clinical research questions into transparent R code while retaining patient-level computation within a local environment. The architecture’s principal contribution is the separation of external LLM-assisted reasoning from local statistical execution, combined with investigator control and preservation of an auditable analytical record. Multidomain validation, formal comparisons with existing analytical approaches, and prospective usability, security, and statistical-validity evaluations are required before broader institutional research deployment.

## Data Availability

This study used data from the Third National Health and Nutrition Examination Survey (NHANES III), a public database conducted by the National Center for Health Statistics (NCHS). The NHANES III protocol was approved by the NCHS Ethics Review Board (ERB). The data are publicly available at https://www.cdc.gov/nchs/nhanes/nhanes3/. Code used in the preparation of this manuscript may be made available upon reasonable request.

## Contributions

S.Y. and Y.H.Y. conceived and designed the study. S.Y. developed the n8n orchestration system and conducted the conversational statistical analyses. S.Z., S.Q., and J.Z. established the retrieval-augmented generation (RAG) framework. W.H.N. and T.Y.H. performed data cleaning and processed the patient-level clinical dataset. V.L.C. and F.J. performed the literature review and curated the bio-statistical guidance validation. Y.H.Y. and F.J. provided administrative, technical, and material support. S.Y. and Y.H.Y. wrote the primary manuscript draft. All authors critically reviewed, revised, and approved the final version of the manuscript.

## Acknowledgments

The study was supported by National Natural Science Foundation of China (82473291), Shaanxi Province “Three Qin Scholars” Innovation Team Project (2023001), and the Fundamental Research Funds for the Central Universities (xtr062023003). The funding body did not play any roles in the design, conduction or reporting of the study.

## Reference

1. Glicksberg, B.S., K.W. Johnson, and J.T. Dudley, The next generation of precision medicine: observational studies, electronic health records, biobanks and continuous monitoring. Hum Mol Genet, 2018. 27(R1): p. R56–r62.

2. Rumsfeld, J.S., K.E. Joynt, and T.M. Maddox, Big data analytics to improve cardiovascular care: promise and challenges. Nat Rev Cardiol, 2016. 13(6): p. 350–9.

3. Atherton, J., Development of the electronic health record. Virtual Mentor, 2011. 13(3): p. 186–9.

4. Rheault, M.N., Optimizing the Electronic Health Record for Clinical Research: Has the Time Come? Kidney360, 2021. 2(12): p. 1880–1881.

5. Sharma, V., et al., Training digitally competent clinicians. Bmj, 2021. 372: p. n757.

6. Team, R.C., RA language and environment for statistical computing, R Foundation for Statistical. Computing, 2020.

7. Delange, B., et al., LinkR: An open source, low-code and collaborative data science platform for healthcare data analysis and visualization. Int J Med Inform, 2025. 199: p. 105876.

8. Clarke, D.J.B., et al., Appyters: Turning Jupyter Notebooks into data-driven web apps. Patterns (N Y), 2021. 2(3): p. 100213.

9. Rathod, V., et al. Privacy and security challenges in large language models. in 2025 IEEE 15th Annual Computing and Communication Workshop and Conference (CCWC). 2025. IEEE.

10. Zhong, X., et al., Considerations for patient privacy of Large Language Models in health care: scoping review. Journal of Medical Internet Research, 2025. 27: p. e76571.

11. Vangala, B.P., et al., Ai-generated code is not reproducible (yet): An empirical study of dependency gaps in llm-based coding agents. arXiv preprint arXiv:2512.22387, 2025.

12. Griot, M., A Methodology for Developing and Integrating Large Language Models into Electronic Health Records to Support Clinical Workflows. 2026, University of Stirling, United Kingdom.

13. Chen, Y., et al., Large Language Models and Prompt-Based Learning: Frontiers and Challenges in Cross-Disciplinary Applications. International Journal of Artificial Intelligence and Robotics Research, 2025. 2: p. 2530001.

14. Collaco, B.G., et al., The role of agentic artificial intelligence in healthcare: a systematic review. 2025.

15. Bo, Y., et al., Development and validation of a prognostic model for MASLD identifying hypertension as a pivotal factor: A population-based study. J Hepatol, 2026. 84(6): p. e206–e208.

16. Dwivedi, S., P. Kasliwal, and S. Soni. Comprehensive study of data analytics tools (RapidMiner, Weka, R tool, Knime). in 2016 Symposium on Colossal Data Analysis and Networking (CDAN). 2016.

17. Devi, M.A., et al. Machine Learning KNIME Based Diseases Prediction for Healthcare Analytics. in 2024 IEEE 21st India Council International Conference (INDICON). 2024.

18. Fillbrunn, A., et al., KNIME for reproducible cross-domain analysis of life science data. J Biotechnol, 2017. 261: p. 149–156.

19. The Galaxy platform for accessible, reproducible, and collaborative data analyses: 2024 update. Nucleic acids research, 2024. 52(W1): p. W83–W94.

20. OpenAI. Introducing Codex CLI. 2025; Available from: https://openai.com/index/introducing-codex/.

21. Anthropic. Claude Code 2025; Available from: https://github.com/anthropics/claude-code.

21. Elucidata. Polly Co-Scientist. 2026; Available from: https://www.elucidata.io/blog/polly-co-scientist.

23. Appsilon. TealFlow. 2025; Available from: https://www.appsilon.com/tealflow.

