## supplemental material for "A Privacy-Preserving Zero-Code Conversational Statistical Analysis System for Clinical Research Using Agentic AI and Local R Execution"

Shujie et al. 2026

#### Appendix A. Detailed Algorithm of Data Structure Perception

This appendix provides the complete logic of the enhanced data structure perception Code node used in Workflow 2. The node is implemented in JavaScript (n8n Code node) and performs three sequential tasks: physical type identification, medical semantic category inference, and distribution feature computation. Missing value detection follows the comprehensive pattern library described in Appendix B.

##### A.1 Physical Type Identification

The algorithm scans up to the first 10,000 rows of the input CSV. For each column, the getDataType(value) function classifies individual values into one of the following categories: null, boolean, date, integer, number, or string. (Arrays or objects are not expected in tabular data; if encountered they are treated as mixed and the column type is resolved accordingly.)

A special pre-check isBinaryBooleanColumn(samples) first tests whether all non-missing values in the column are binary – i.e., 0/1, true/false, or any of the Boolean string equivalents defined below. If this check passes, the column is directly assigned the boolean physical type without further promotion.

For columns that are not purely binary, each non-missing value is processed by promoteType(current, new), which follows a strict precedence order:

null < boolean < date < integer < number < string < mixed.

For example, a column containing both integers (e.g., 5) and floats (e.g., 5.2) will be resolved as number. The precedence ensures that the most informative type is retained.

Boolean string equivalents (case-insensitive matching) are recognised by isBooleanString(value):

True equivalents: "true", "yes", "1", "on", "y", "t", "是", "真", "ok", "enable", "enabled", "active", "activated".

False equivalents: "false", "no", "0", "off", "n", "f", "否", "假", "cancel", "disable", "disabled", "inactive", "deactivated".

Date detection is performed by isDateString(value). The function applies four regular expression patterns that cover common date formats: YYYY-MM-DD, DD/MM/YYYY, ISO 8601 (with optional time and timezone), and DD Mon YYYY. Additionally, it verifies that parsing the string with new Date() yields a year between 1900 and 2100, while discarding plain numeric strings that might be misinterpreted as dates.

**A.2 Medical Semantic Category Inference**

After physical typing, the function getMedicalCategory(name, type, samples, totalRows) assigns each column one of five semantic categories: continuous, categorical, ordinal, identifier, or date_time. The inference follows a strict priority hierarchy, with a newly introduced suffix override rule for biomarker-named columns that clearly indicate ordinal meaning.

Biomarker protection – If isClinicalBiomarker(name) returns true, the column is normally classified as continuous regardless of its physical type or uniqueness. This prevents critical laboratory measurements (e.g., hdl, creatinine, glucose) from being misidentified as identifiers or categorical variables. The biomarker library includes a comprehensive set of keywords (e.g., alt, ast, hdl, ldl, creatinine, glucose, hba1c, crp, etc.) as well as pattern-based matches such as plasma_glucose, hdl_cholesterol, total_cholesterol. Matching is case-insensitive and ignores underscores (underscores are treated as separators, so hdl_cholesterol is matched by the presence of hdl).

Suffix override rule – If a column passes the biomarker test and its name ends with one of the following ordinal suffixes: _grade, _stage, _class, _level, _severity, _intensity, _score, _rank, the algorithm then calls isLikelyOrdinalVariable(name, samples, type). If the ordinal check succeeds (i.e., the column is considered ordinal based on its values), the column is re-classified as ordinal instead of continuous. This rule ensures that derived variables such as hdl_grade or stage_biomarker are correctly treated as ordered categories when their actual values match ordinal patterns, while truly continuous biomarker measurements (e.g., hdl_value) remain continuous.

Identifier detection – isIdentifierColumn(name, type, samples, total, uniq) runs only for columns that are not protected as biomarkers. It checks the column name against a keyword library using regular expressions: ^id$, ^subject_id$, ^patient_id$, ^participant_id$, ^seqn$, ^record_id$. A column is marked as identifier if it passes the keyword test and either (a) the ratio of unique values among valid samples exceeds 0.99, or (b) the number of distinct values equals the total row count. An additional safeguard excludes columns with very low coefficient of variation (indirectly) and, more importantly, ensures that any column already flagged as a biomarker is never considered an identifier.

Ordinal variable detection – The function isLikelyOrdinalVariable(name, samples, type) first checks if the column contains ordinal keywords (e.g., stage, grade, level, degree, class, phase, severity, intensity, score, rank). It then validates whether the actual values conform to any of the following specific medical scale patterns. For each scale, the detection requires that the column name includes at least one associated keyword and that every non-missing value in the sample matches the corresponding regular expression. The implemented scales are:

ECOG / Zubrod / performance status – values 0–4.

Child-Pugh – values A, B, C (case-insensitive).

NYHA – values I, II, III, IV.

Glasgow Coma Scale (GCS) – values 3–15.

Karnofsky Performance Status (KPS) – values 0, 10, 20, 30, 40, 50, 60, 70, 80, 90, 100.

Modified Rankin Scale (mRS) – values 0–6.

SOFA (Sequential Organ Failure Assessment) – values 0–24.

qSOFA (quick SOFA) – values 0–3.

ASA Physical Status – values 1–6.

CTCAE Grade (Common Terminology Criteria for Adverse Events) – values 1–5.

If any of these scale patterns match, the column is considered ordinal and the specific scale name is recorded (e.g., karnofsky, mrs, sofa). Otherwise, a generic ordinal classification is assigned if an ordinal keyword is present and the number of unique values is between 2 and 10 (inclusive). Columns that fail all ordinal criteria proceed to the remaining mapping.

Remaining type mapping – After the priority steps (biomarker, identifier, ordinal), the decision proceeds as follows:

Physical type number → continuous.

Physical type date → date_time.

Physical type boolean → categorical.

Physical type integer → categorical if the number of unique values is ≤ 10 or the column name contains score or grade; otherwise continuous.

For all other cases (e.g., low-cardinality strings that are not captured by earlier rules) – ordinal if the number of unique values is ≤ 5, else categorical.

This hierarchy ensures that clinically meaningful categories are prioritised and that rare or ambiguous cases fall back to sensible defaults.

**A.3 Distribution Feature Calculation and Normality Assessment**

For each column, the node computes a rich set of distribution features that are subsequently used in the workflow’s quality report and data profiling summary.

For categorical, ordinal, and Boolean columns – the following statistics are extracted:

Missing rate (proportion of missing values in the scanned sample).

Number of unique (distinct) non-missing values.

Frequency table of the top five most common values, each with its absolute count and proportion (rounded to three decimals).

A rare category flag – set to true if any category has an absolute count < 5.

A low-variability flag – set to true if the most frequent category accounts for more than 95% of non-missing entries.

For continuous columns (excluding columns with physical type string or date) – the following features are computed:

Missing rate.

Mean, standard deviation, minimum, and maximum – each rounded to three decimal places.

A five-bin equal-width frequency distribution. The bin boundaries are determined from the observed minimum and maximum values, and each bin reports its range and the proportion of non-missing values falling into that bin.

A comprehensive normality assessment via assessNormality(nums) (see below).

Normality assessment – For a numeric array, the algorithm first checks the sample size. If $n < 8$, the verdict is "N/A" (insufficient data). For $n \geq 8$, it calculates the skewness and excess kurtosis using moment-based estimators:

$$\text{Skewness} = (1/n)\, \Sigma((x_i - \mu)^3) \,/\, (\, (1/n)\, \Sigma((x_i - \mu)^2)\, )^{3/2}$$

$$\text{Excess kurtosis} = (1/n)\, \Sigma((x_i - \mu)^4) \,/\, (\, (1/n)\, \Sigma((x_i - \mu)^2)\, )^2 - 3$$

A column is considered Normal if $|\text{skewness}| < 1.5$ and $|\text{kurtosis}| < 3$; otherwise it is flagged as Non-Normal. The output includes the verdict, the skewness value, and the kurtosis value (both rounded to three decimals). This rule-based approach is robust for moderate to large sample sizes and avoids the computational overhead of formal hypothesis tests when not strictly required.

Rare variable flag (isR) – A column is flagged as potentially “rare” (highly sparse or invariant) if:

Its missing rate exceeds 0.9, or

It is categorical, ordinal, or Boolean and either rare categories exist (any category count < 5) or the distribution shows low variability (dominant category > 95%).

This flag helps downstream processes (such as feature selection or anomaly detection) to identify columns that may contribute little information.

All distribution features are stored per column as part of the output JSON, enabling further analysis or visualisation in subsequent workflow nodes.

#### Appendix B. Missing Value Handling Details

The following lists the exact strings recognized as missing values by the Code node. The detection is case-sensitive for some patterns and case-insensitive for others, as implemented in the workflow.

Exact string matches (case- sensitive, unless noted):

'' (empty string), 'null', 'undefined', 'nan', 'none', 'na', 'nil', 'NULL', 'UNDEFINED', 'NAN', 'NONE', 'NA', 'NIL', 'Null', 'Undefined', 'Nan', 'None', 'Na', 'Nil', 'N/A', 'n/a', '#N/A', '#NA', '#NULL', '#VALUE!', '#REF!', '#DIV/0!', '#NAME?', 'missing', 'MISSING', 'Missing', 'absent', 'ABSENT', 'Absent', 'NotDone', 'void', 'VOID', 'Void', 'empty', 'EMPTY', 'Empty', 'unknown', 'UNKNOWN', 'Unknown', 'not available', 'NOT AVAILABLE', 'Not Available', '\\N', '(null)', '(NULL)', '(undefined)', '(none)', '-', '--', '---', '.', '..', '...', '-1', '-999', '-9999', '(space)', ' (fullwidth space)', '\t', '\n', '\r',

Chinese equivalents: '空', '空值', '缺失', '无', '未知', '未填写', '未提供', '不适用',

Spanish/French: 'vacío', 'VACÍO', 'nulo', 'NULO', 'manquant', 'MANQUANT'.

Regular expressions for fuzzy matching (applied after trimming):

/^\s*$/ (whitespace only),

/^null$/i, /^nan$/i, /^none$/i, /^na$/i, /^n\/a$/i,

/^undefined$/i, /^missing$/i, /^#n\/?a$/i,

/^\?+$/ (one or more question marks),

/^-+$/ (one or more hyphens),

/^\.+$/ (one or more dots),

/^*+$/ (one or more asterisks),

/^\\n$/i, /^\(null\)$/i, /^\(none\)$/i.

### Appendix C. Outlier Detection Formulas and Thresholds

#### C.1 Detection Methods and Outliers Report Generation

Both detection methods are applied simultaneously to every column classified as continuous. The default parameters are:

Interquartile Range (IQR) Method:

IQR = Q3 – Q1

Lower fence = Q1 – 1.5 × IQR

Upper fence = Q3 + 1.5 × IQR

Values below the lower fence or above the upper fence are flagged as outliers.

Z- score Method:

Z = (x – mean) / standard deviation

Values with |Z| > 3 are flagged as outliers.

The multiplier for the IQR method (1.5) and the Z-score threshold (3) are default values. In our workflow, the researcher can modify these parameters at any time through the chat interface. For example, they may specify a 2.5× IQR rule or a |Z| > 2.5 criterion, depending on the characteristics of the dataset or domain requirements. This flexibility allows the outlier detection strategy to be tailored without altering the underlying code.

Output Format:

Outlier.csv: a table with columns variable, method (IQR / Z- score), outlier_count, min_outlier, max_outlier, and all outliers (comma- separated, descending).

Outlier.txt: a human- readable text report summarizing the above information with clear labels for each detected column.

#### C.2 Outlier Treatment Input Specification

In our workflow, after the outlier detection report is generated, the researcher is prompted to specify the treatment strategy for the identified outliers. Rather than relying on free-text descriptions, which may introduce ambiguity and parsing errors, we provide a structured command template directly within the chat interface. This template serves as a reference guide, informing the researcher of the available operation types (Remove, Cap, Replace with NA, and No action) and the expected syntax for combining methods, target columns, and optional remarks.

The researcher reads the outlier report and then formulates a response following the provided template (e.g., "Using the 1.5× IQR rule, remove outliers for age, cap outliers for BUN, and take no action for other variables."). This approach ensures that the outlier-cleaning strategy is clearly documented, reproducible, and machine-parseable without requiring manual interpretation of free-form text. It also allows the researcher to easily adjust the IQR multiplier (e.g., 2.5×) or add variable-specific instructions as needed. If no outlier treatment is desired, the researcher may simply reply with the predefined phrase indicating that the original data will be passed through unchanged.

The full input instructions, including the operation definitions and usage hints, are reproduced below.

Please enter your final outlier analysis requirements.

Operation Instructions

"Remove": Remove the entire row containing outliers (filter).

"Cap": Replace values outside the [lower bound, upper bound] with the nearest bound value (lower or upper).

"Replace with NA": Set outliers to NA.

"No action": Keep original values, no outlier detection or modification performed.

Command Template (Method | Target Column | Action Verb | Remarks)

Using the 1.5× IQR rule, remove outliers for age, cap outliers for BUN, and take no action for other variables.

Usage Hints

Strictly follow the verbs and target columns above. Do not change the method on your own.

Replace the column names and verbs in the template with those of your actual dataset.

If all columns require "no action", simply reply: "No outlier treatment will be performed on the data. The original file will be read and saved as a CSV file only."

If you need to change the IQR multiplier (e.g., 2.5×), specify it clearly.

Please strictly follow the above format.

### Appendix D. Pinecone Knowledge Base Full Module Description

R Programming Standards Module: naming conventions, code structure templates.

The Pinecone index (n8ntest) stores vector embeddings of five categories of domain knowledge. Documents are chunked with a size of 2,500 characters and an overlap of 500 characters before embedding with the OpenAI or Google Gemini embedding model.

**Module 1: R Programming Standards**

Contains best-practice guidelines for R code style, variable naming conventions (e.g., snake_case), function documentation with # comments, and recommended package loading sequences.

**Module 2: Publication‑Grade Figure Template Library**

Pre‑written ggplot2 templates optimised for major medical journals. Parameters include:

Colour palettes: scale_color_brewer(), scale_fill_viridis_d().

Theme: theme_minimal() with text = element_text(family = "Times New Roman", size = 12).

Resolution: ggsave(..., dpi = 300, width = 8, height = 6, units = "in").

Specific templates exist for boxplots, Kaplan‑Meier curves, forest plots, heatmaps, and nomograms.

**Module 3: Clinical Statistical Strategy Library**

Validated logic blocks for common analytical scenarios:

Group comparison: two‑sample t‑test (normal), Mann‑Whitney U (non‑normal), chi‑squared/Fisher's exact test (categorical).

Survival analysis: Cox proportional hazards model with proportionality assumption check, Kaplan‑Meier estimation with log‑rank test.

Regression: linear, logistic, and ordinal logistic regression with variable selection strategies.

Diagnostic performance: ROC curve with pROC, sensitivity/specificity.

**Module 4: Medical Research R Package Navigation Library**

Maps research tasks to recommended packages:

Descriptive tables → gtsummary

Survival analysis → survival, survminer

Competitive risk → cmprsk

Machine learning → caret, tidymodels, glmnet

Table formatting → kableExtra, openxlsx

**Module 5: Interactive Requirement Template (SOP) Library**

Structured parameter checklists for complex visualizations. For example, a heatmap template asks: “Should rows/columns be clustered? Distance metric? Clustering method? Should the colour scale be standardized by row? What colour palette?” A KM- curve template asks: “Which variable defines groups? Should the risk table be displayed? Should confidence intervals be shown? At what time point should the curve be truncated?”

### Appendix E. Supplementary Results

In this section, we present the full analytical pipeline executed on our workflow tool, together with all resultant analytical figures and tables.All analyses, except those presented in Section E.2, were performed in Workflow 2.

#### E.1 Complete workflow demonstration

The complete, end-to-end operational logic of the n8n orchestration pipeline is visually detailed in Figure S1.

*Figure S1. Native node configuration and data-flow pathways of the n8n orchestration backend.* ***a, b*** *Workflow 1: Automated Data Preprocessing.* ***c*** *Workflow 2: Automated Statistical Analysis & Reporting.*

a

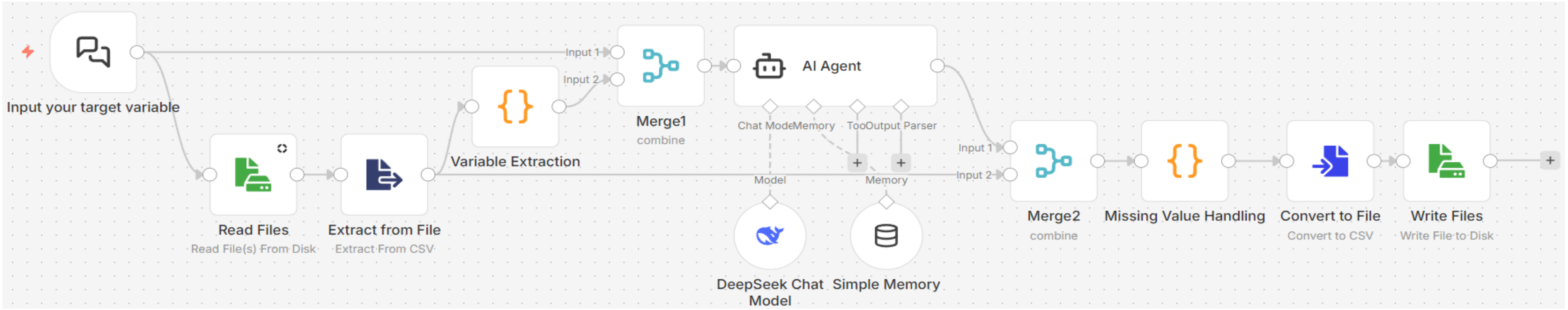


b

c

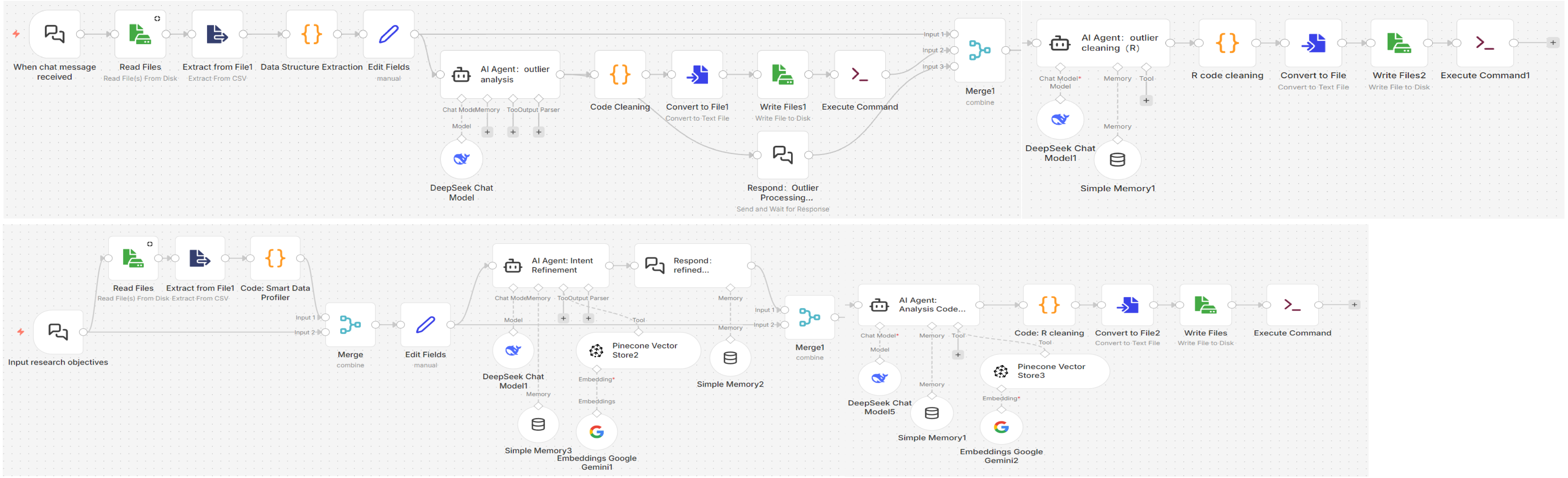

**E.2 Missing Value Removal and outlier processing**

Chat Trigger: Define the following as target variables: sex, age, race-ethnicity, race, ethnicity, drinkever, total_bilirubin, total_protein, mortstat, permth_exm, permth_int, liver_ultrasound, serum_insulin, alanine_aminotransferase, creatinine, blood_urea_nitrogen, alkaline_phosphatase, aspartate_aminotransferase, gamma_glutamyl_transferase, platelet, fasting_blood_glucose, bmi, waist_circumference, sbp, dbp, total_cholesterol, serum_triglycerides, and hdl_cholesterol.

Response to Chat: No outliers with clinical significance were detected among the variables included in the analysis; therefore, no outlier processing will be conducted, and the original dataset shall be exported directly.

**E.3 Kaplan-Meier Curve Generation**

Chat Trigger: "Please generate Kaplan-Meier curves for patients with metabolic liver disease, stratified by the presence or absence of hypertension. Display key time points at 0, 100, 200, 300, and 400 months."

Response to Chat: "Please generate Kaplan-Meier curves for patients with metabolic liver disease, stratified by the presence or absence of hypertension, with key time points displayed at 0, 100, 200, 300, and 400 months. Use 'mortstat' as the event variable and 'permth_int' (follow-up time in months) as the survival time variable. Metabolic dysfunction-associated steatotic liver disease(MASLD) is defined by a `maf_5` ≥ 0. Hypertension is defined according to clinical guidelines (i.e., `sbp` ≥ 140 mmHg and/or `dbp` ≥ 90 mmHg)."

***Figure S2. Kaplan-Meier survival curves for all-cause mortality stratified by hypertension status in MASLD patients***

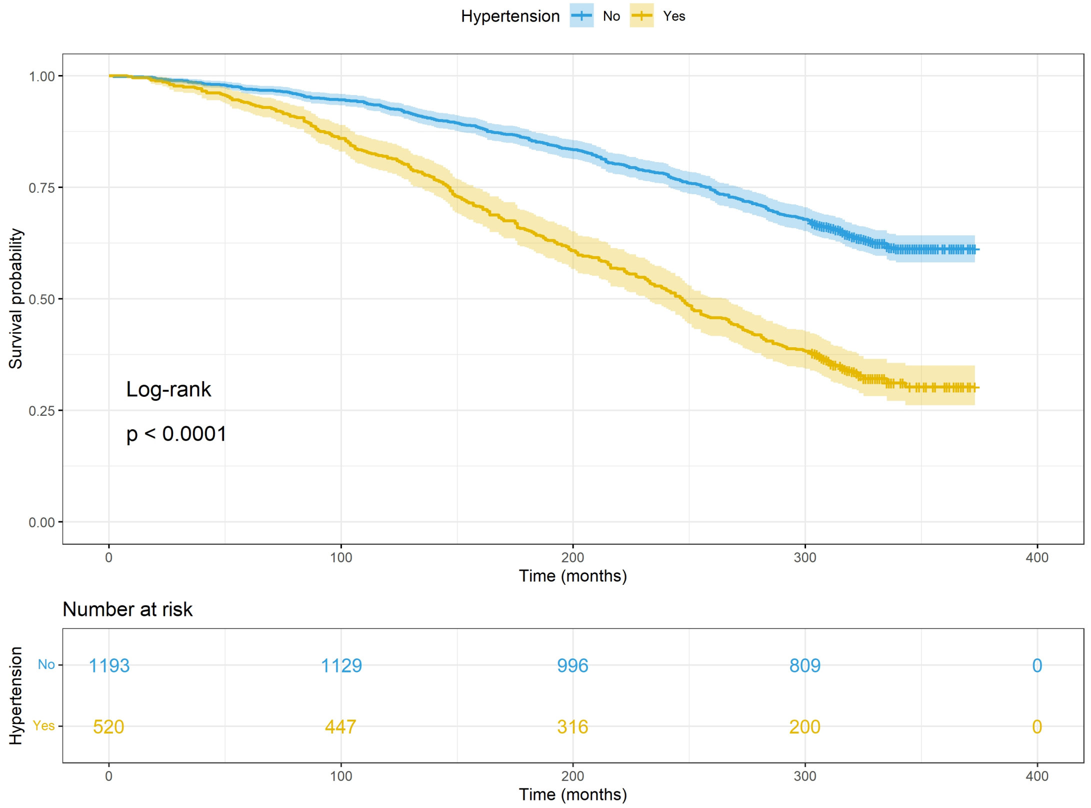


Kaplan-Meier curves showing the overall survival probability of MASLD patients with (yellow, Yes) and without (blue, No) hypertension, with shaded areas indicating 95% confidence intervals for survival estimates. The log-rank test demonstrated a significant between-group difference in survival outcomes ($p<0.0001$). The below risk table presents the number of participants remaining at risk at each prespecified time point (0, 100, 200, 300, 400 months) for the two subgroups respectively.

##### E.4 Cox Regression (Variable Selection Using Univariate Cox Regression and Stepwise Regression)

###### *Descriptive Statistical Analysis and Univariate Cox Regression*

Chat trigger: Univariate Cox regression analysis was conducted using `mortstat` as the event variable (1 indicating death) and `permth_int` (follow-up time) as the survival time variable. The following variables were individually analyzed for their association with mortality: age, sex, race-ethnicity, race, ethnicity, bmi, waist_circumference, sbp, dbp, total_cholesterol, serum_triglycerides, hdl_cholesterol, fasting_blood_glucose, aspartate_aminotransferase, alanine_aminotransferase, alkaline_phosphatase, gamma_glutamyl_transferase, platelet, serum_insulin, total_bilirubin, total_protein, crp, creatinine, blood_urea_nitrogen, diabetes, hba1c.

Respond to chat: Univariate Cox regression analysis was conducted using `mortstat` as the event variable (1 indicating death) and `permth_int` (follow-up time) as the survival time variable. The following variables were individually analyzed for their association with mortality:age, sex, race-ethnicity, race, ethnicity, bmi, waist_circumference, sbp, dbp, total_cholesterol, serum_triglycerides, hdl_cholesterol, fasting_blood_glucose, aspartate_aminotransferase, alanine_aminotransferase, alkaline_phosphatase, gamma_glutamyl_transferase, platelet, serum_insulin, total_bilirubin, total_protein, crp, creatinine, blood_urea_nitrogen, diabetes, hba1c. Descriptive statistics were presented stratified by survival status (`mortstat` = 0 vs. 1). Continuous variables were reported as median (interquartile range), and categorical variables were reported as frequency (percentage). The proportional hazards assumption (linearity assumption) for continuous variables was assessed using Cox regression residual-based methods, and variables identified as violating this assumption were labeled "nonlinear variable."

The results were compiled into tables containing the following information for each variable: variable name, category level, hazard ratio (HR), 95% confidence interval (lower and upper limits), and *p*-value (with significance markers: * $p < 0.05$, ** $p < 0.01$, *** $p < 0.001$). Nonlinear variables were annotated accordingly.

***Table S1. Baseline clinical characteristics of MAFLD patients stratified by all-cause mortality status.***

| Variable | Overall<br>N = 7,942[1] | Alive<br>N = 5,688[1] | Died<br>N = 2,254[1] | ***p*-value**[2] |
|---|---|---|---|---|
| **age** | 39.00 (29.00, 54.00) | 34.00 (27.00, 43.00) | 60.00 (47.00, 68.00) | <0.001 |
| **bmi** | 26.40 (23.20, 30.30) | 26.00 (22.90, 30.00) | 27.40 (24.10, 31.10) | <0.001 |
| **waist_circumference** | 91.70 (81.60, 101.80) | 89.30 (79.30, 99.10) | 97.30 (88.30, 106.40) | <0.001 |
| **sbp** | 119.00 (110.00, 130.00) | 115.00 (108.00, 124.00) | 131.00 (119.00, 146.00) | <0.001 |
| **dbp** | 74.00 (67.00, 81.00) | 73.00 (66.00, 79.00) | 77.00 (70.00, 84.00) | <0.001 |
| **total_cholesterol** | 198.00 (171.00, 228.00) | 192.00 (167.00, 220.00) | 213.00 (185.00, 243.00) | <0.001 |
| **serum_triglycerides** | 107.00 (76.00, 162.00) | 99.50 (71.00, 148.00) | 133.00 (90.00, 189.00) | <0.001 |
| **hdl_cholesterol** | 48.00 (40.00, 58.00) | 49.00 (41.00, 58.00) | 47.00 (39.00, 58.00) | <0.001 |
| **fasting_blood_glucose** | 92.60 (86.70, 99.90) | 91.10 (85.90, 97.50) | 97.20 (90.30, 108.70) | <0.001 |
| **aspartate_aminotransferase** | 19.00 (16.00, 24.00) | 19.00 (16.00, 23.00) | 19.00 (16.00, 24.00) | <0.001 |
| **alanine_aminotransferase** | 14.00 (10.00, 21.00) | 15.00 (10.00, 21.00) | 14.00 (10.00, 20.00) | 0.002 |
| **alkaline_phosphatase** | 81.00 (67.00, 98.00) | 78.00 (64.00, 95.00) | 88.00 (72.00, 106.00) | <0.001 |
| **gamma_glutamyl_transferase** | 22.00 (15.00, 33.00) | 20.00 (14.00, 31.00) | 24.00 (17.00, 39.00) | <0.001 |
| **platelet** | 265.50 (226.50, 311.00) | 267.00 (228.50, 312.00) | 260.50 (221.00, 307.50) | <0.001 |
| **serum_insulin** | 54.96 (38.28, 83.88) | 52.80 (37.26, 79.20) | 60.78 (41.88, 97.80) | <0.001 |
| **total_bilirubin** | 0.50 (0.40, 0.70) | 0.50 (0.40, 0.70) | 0.50 (0.40, 0.70) | 0.2 |
| **total_protein** | 7.40 (7.10, 7.70) | 7.40 (7.10, 7.70) | 7.30 (7.00, 7.70) | <0.001 |
| **crp** | 0.21 (0.21, 0.44) | 0.21 (0.21, 0.33) | 0.21 (0.21, 0.60) | <0.001 |
| **creatinine** | 1.00 (0.90, 1.20) | 1.00 (0.90, 1.10) | 1.10 (0.90, 1.20) | <0.001 |

| **Variable** | **Overall**<br>N = 7,942[1] | **Alive**<br>N = 5,688[1] | **Died**<br>N = 2,254[1] | ***p*-value**[2] |
|---|---|---|---|---|
| **blood_urea_nitrogen** | 13.00 (10.00, 16.00) | 12.00 (10.00, 15.00) | 14.00 (11.00, 17.00) | <0.001 |
| **hba1c** | 5.30 (5.00, 5.70) | 5.20 (4.90, 5.50) | 5.60 (5.20, 6.10) | <0.001 |
| **sex** | | | | <0.001 |
| Female | 4,448 (56.0%) | 3,312 (58.2%) | 1,136 (50.4%) | |
| Male | 3,494 (44.0%) | 2,376 (41.8%) | 1,118 (49.6%) | |
| **race_ethnicity** | | | | <0.001 |
| Mexicanamerican | 2,232 (28.1%) | 1,754 (30.8%) | 478 (21.2%) | |
| Nonhispanicwhite | 5,312 (66.9%) | 3,621 (63.7%) | 1,691 (75.0%) | |
| Other | 398 (5.0%) | 313 (5.5%) | 85 (3.8%) | |
| **race** | | | | <0.001 |
| Black | 2,525 (31.8%) | 1,801 (31.7%) | 724 (32.1%) | |
| Mexican_American_unknown_race | 3 (0.0%) | 2 (0.0%) | 1 (0.0%) | |
| Other | 357 (4.5%) | 291 (5.1%) | 66 (2.9%) | |
| White | 5,057 (63.7%) | 3,594 (63.2%) | 1,463 (64.9%) | |
| **ethnicity** | | | | <0.001 |
| Mexican_American | 2,232 (28.1%) | 1,754 (30.8%) | 478 (21.2%) | |
| Not_Hispanic | 5,455 (68.7%) | 3,735 (65.7%) | 1,720 (76.3%) | |
| Other_Hispanic | 255 (3.2%) | 199 (3.5%) | 56 (2.5%) | |
| **diabetes** | | | | <0.001 |
| 1 | 492 (6.2%) | 175 (3.1%) | 317 (14.1%) | |

| Variable | Overall<br>N = 7,942[1] | Alive<br>N = 5,688[1] | Died<br>N = 2,254[1] | *p*-value[2] |
|---|---|---|---|---|
| 2 | 7,450 (93.8%) | 5,513 (96.9%) | 1,937 (85.9%) | |

[1]Continuous variables are expressed as median (IQR), and categorical variables are presented as n (%).

[2]Wilcoxon rank sum test; Fisher's exact test; Fisher's Exact Test for Count Data with simulated p-value. (based on 2000 replicates)

Abbreviations: bmi, body mass index; sbp, systolic blood pressure; dbp, diastolic blood pressure; hdl, high-density lipoprotein; crp, C-reactive protein; HbA1c, glycated hemoglobin.

***Table S2. Univariate Cox proportional hazards regression analysis for all-cause mortality among patients with MASLD***

| Variable | HR | 95% CI | *p*-value |
|---|---|---|---|
| **age** | 1.09 | 1.08, 1.09 | <0.001 |
| **bmi** | 1.03 | 1.02, 1.04 | <0.001 |
| **waist_circumference** | 1.03 | 1.03, 1.03 | <0.001 |
| **sbp** | 1.01 | 1.01, 1.01 | <0.001 |
| **dbp** | 1.00 | 1.00, 1.00 | <0.001 |
| **total_cholesterol** | 1.01 | 1.01, 1.01 | <0.001 |
| **serum_triglycerides** | 1.00 | 1.00, 1.00 | <0.001 |
| **hdl_cholesterol** | 1.00 | 1.00, 1.00 | 0.486 |
| **fasting_blood_glucose** | 1.01 | 1.01, 1.01 | <0.001 |
| **aspartate_aminotransferase** | 1.00 | 1.00, 1.01 | <0.001 |
| **alanine_aminotransferase** | 1.00 | 0.99, 1.00 | 0.01 |
| **alkaline_phosphatase** | 1.01 | 1.00, 1.01 | <0.001 |
| **gamma_glutamyl_transferase** | 1.00 | 1.00, 1.00 | <0.001 |
| **platelet** | 1.00 | 1.00, 1.00 | <0.001 |
| **serum_insulin** | 1.00 | 1.00, 1.00 | 0.753 |
| **total_bilirubin** | 0.97 | 0.85, 1.10 | 0.616 |
| **total_protein** | 0.85 | 0.78, 0.94 | <0.001 |
| **crp** | 1.00 | 1.00, 1.00 | 0.633 |
| **creatinine** | 1.00 | 1.00, 1.00 | 0.018 |
| **blood_urea_nitrogen** | 1.06 | 1.05, 1.06 | <0.001 |
| **hba1c** | 1.00 | 1.00, 1.00 | 0.384 |
| **sex** | | | |
| Female | — | — | |
| Male | 1.30 | 1.20, 1.42 | <0.001 |
| **race_ethnicity** | | | |
| Mexicanamerican | — | — | |
| Nonhispanicwhite | 1.55 | 1.40, 1.72 | <0.001 |
| Other | 0.96 | 0.76, 1.21 | 0.74 |
| **race** | | | |
| Black | — | — | |
| Mexican_American_unknown_race | 1.38 | 0.19, 9.80 | 0.748 |
| Other | 0.61 | 0.47, 0.78 | <0.001 |
| White | 1.01 | 0.92, 1.10 | 0.898 |
| **ethnicity** | | | |
| Mexican_American | — | — | |
| Not_Hispanic | 1.53 | 1.38, 1.69 | <0.001 |
| Other_Hispanic | 0.99 | 0.75, 1.31 | 0.951 |
| **diabetes** | | | |
| 1 | — | — | |
| 2 | 0.28 | 0.25, 0.32 | <0.001 |

All analyses were unadjusted. Variables marked with an em dash (—) served as the reference group for categorical covariates. Abbreviations: HR, hazard ratio; CI, confidence interval.

***Stepwise Regression for Variable Selection and Model Construction***

Chat trigger: Mortstat was used as the event variable for mortality (1 = death), and permth_int (follow-up time, in months) was used as survival time. MASLD, defined by `maf_5` ≥ 0, was used as the study condition. Within the MASLD population, stepwise regression was performed by incorporating the following variables to select and finalize the variable list for the final model: age, sex, race-ethnicity, bmi, waist_circumference, sbp, dbp, total_cholesterol, serum_triglycerides, fasting_blood_glucose, aspartate_aminotransferase, alanine_aminotransferase, alkaline_phosphatase, gamma_glutamyl_transferase, platelet, total_bilirubin, total_protein, creatinine, blood_urea_nitrogen, diabetes.

Respond to chat: Mortstat was used as the event variable for mortality (1 = death), and permth_int (follow-up time, in months) was used as survival time. MASLD, defined by `maf_5` ≥ 0, was used as the study condition. Within the MASLD population, stepwise regression was performed by incorporating the following variables to select and finalize the variable list for the final model: age, sex, race-ethnicity, bmi, waist_circumference, sbp, dbp, total_cholesterol, serum_triglycerides, Fasting_blood_glucose, aspartate_aminotransferase, alanine_aminotransferase, alkaline_phosphatase, gamma_glutamyl_transferase, platelet, total_bilirubin, total_protein, creatinine, blood_urea_nitrogen, diabetes. The following outputs were required: the final list of included variables along with their coefficients, hazard ratios (HRs), 95% confidence intervals (CIs), and *p*-values; overall model performance metrics, including the concordance index (C-index) and its 95% CI, Akaike information criterion (AIC), and Bayesian information criterion (BIC); time-dependent receiver operating characteristic (ROC) curves with corresponding area under the curve (AUC) values for 5-, 10-, and 20-year time points (generated using the timeROC package and combined into a single figure); and calibration curves. The stepwise regression algorithm employed a backward selection method with *p*-values as the criterion for variable entry and removal, using a p-out threshold of 0.05. Convergence was defined as two consecutive iterations with no variable changes. Continuous variables were included in their original, untransformed form. The proportional hazards assumption was evaluated by performing Schoenfeld residual tests on the variables included in the final model. Multicollinearity was assessed by calculating the variance inflation factor (VIF) for continuous variables in the final model.

Based on the above dialogue, we constructed a multivariate Cox proportional hazards regression model via stepwise regression using the analysis workflow embedded in our n8n tool to identify independent predictors of all-cause mortality among patients with MASLD (Table S3). Advanced age (HR=1.056, 95% CI: 1.049–1.064, P<0.0001), larger waist circumference (HR=1.017, 95% CI: 1.005–1.028, P=0.004), elevated systolic blood pressure (HR=1.014, 95% CI: 1.010–1.018, P<0.0001), higher fasting blood glucose (HR=1.003, 95% CI: 1.002–1.004, P<0.0001), elevated aspartate aminotransferase (HR=1.007, 95% CI: 1.004–1.011, P<0.0001), elevated gamma-glutamyl transferase (HR=1.003, 95% CI: 1.002–1.003, P<0.0001), higher total bilirubin (HR=1.302, 95% CI: 1.083–1.565, P=0.005), and increased serum creatinine (HR=1.279, 95% CI: 1.132–1.446, P<0.0001) were significantly associated with an elevated risk of all-cause mortality in MASLD patients.

In contrast, higher body mass index (BMI) (HR=0.973, 95% CI: 0.949–0.996, P=0.025), elevated diastolic blood pressure (HR=0.988, 95% CI: 0.984–0.992, P<0.0001), higher total cholesterol (HR=0.998, 95% CI: 0.996–0.999, P=0.034), elevated alanine aminotransferase (HR=0.987, 95% CI: 0.982–0.993, P<0.0001), and higher platelet count (HR=0.998, 95% CI: 0.997–0.999, P=0.013) served as independent protective factors against all-cause mortality for patients with MASLD.

***Table S3. Multivariate Cox proportional hazards regression model for all-cause mortality among patients with MASLD.***

| Variable | HR | 95% CI Lower | 95% CI Upper | *p*-value |
|---|---|---|---|---|
| **age** | 1.056 | 1.049 | 1.064 | <0.0001 |
| **bmi** | 0.973 | 0.949 | 0.996 | 0.025 |
| **waist_circumference** | 1.017 | 1.005 | 1.028 | 0.004 |
| **sbp** | 1.014 | 1.01 | 1.018 | <0.0001 |
| **dbp** | 0.988 | 0.984 | 0.992 | <0.0001 |
| **total_cholesterol** | 0.998 | 0.996 | 0.999 | 0.034 |
| **fasting_blood_glucose** | 1.003 | 1.002 | 1.004 | <0.0001 |
| **aspartate_aminotransferase** | 1.007 | 1.004 | 1.011 | <0.0001 |
| **alanine_aminotransferase** | 0.987 | 0.982 | 0.993 | <0.0001 |
| **gamma_glutamyl_transferase** | 1.003 | 1.002 | 1.003 | <0.0001 |
| **platelet** | 0.998 | 0.997 | 0.999 | 0.013 |
| **total_bilirubin** | 1.302 | 1.083 | 1.565 | 0.005 |
| **creatinine** | 1.279 | 1.132 | 1.446 | <0.0001 |

Abbreviations: HR, hazard ratio; 95% CI, 95% confidence interval. Variables retained in the final multivariate Cox regression model were screened by stepwise regression. Hazard ratios greater than 1 indicate increased risk of all-cause mortality, while HR values less than 1 represent reduced mortality risk.

***Table S4. Predictive Performance and Goodness-of-Fit Diagnostics of the Prognostic Model***

| Metric | Value |
|---|---|
| **C-index** | 0.7781 |
| **C-index_lower** | 0.7617 |
| **C-index_upper** | 0.7944 |
| **AIC** | 10471.5891 |
| **BIC** | 10532.3420 |

The table metrics present the statistical validation diagnostics evaluated for the established prognostic regression model. Model discrimination capacity is quantified using Harrell's Concordance Index (C-index), where a value of 0.7781 indicates a robust predictive accuracy for the time-to-event outcomes, bounded by a 95% confidence interval spanning from 0.7617 to 0.7945. Model parsimony, calibration quality, and relative information loss are parsed simultaneously through the Akaike Information Criterion (AIC = 10471.5891) and the Bayesian Information Criterion (BIC = 10532.3420). These cumulative diagnostic fingerprints serve as baseline metrics for downstream model comparison and cross-validation pipelines.

Abbreviations: AIC, Akaike Information Criterion; BIC, Bayesian Information Criterion; C-index, Concordance Index.

##### E.5 Forest Plot Generation

Chat trigger: A multivariable Cox regression analysis was conducted to identify independent risk factors for mortality in patients with metabolic dysfunction-associated steatotic liver disease (MASLD). Mortality status was defined using the variable `mortstat` (1 = death), and survival time was measured using `permth_int` (follow-up time in months). MASLD was defined based on the variable `maf_5` ≥ 0. The following variables were directly included in the regression model: age, bmi, waist_circumference, sbp, dbp, total_cholesterol, fasting_blood_glucose, aspartate_aminotransferase, platelet, alanine_aminotransferase, gamma_glutamyl_transferase, total_bilirubin, creatinine.

Respond to chat: Prior to regression analysis, the proportional hazards assumption was tested for all independent variables. Variables that violated this assumption were reported. The model used `mortstat` as the event variable and `permth_int` as the survival time. The study population consisted of patients with MASLD defined by `maf_5` ≥ 0. A forest plot was generated with the following specifications: the right side of the plot displays the variable names, hazard ratios (HR) with 95% confidence intervals (CI), and *p*-values; the left side presents the graphical representation of the forest plot, including the point estimate of the HR (depicted as a square) and the 95% CI (depicted as a line segment) for each variable. A vertical dashed reference line was added at x = 1.0 (indicating HR = 1, representing no effect). The following variables were directly included:age, bmi, waist_circumference, sbp, dbp, total_cholesterol, fasting_blood_glucose, aspartate_aminotransferase, platelet, alanine_aminotransferase, gamma_glutamyl_transferase, total_bilirubin, creatinine.

***Figure S3. Forest plot of independent predictors for all-cause mortality among MASLD patients.***

| Variable | HR (95% CI) | P Value |
|---|---|---|
| Age | 1.06 (1.05−1.06) | <0.001 |
| BMI | 0.97 (0.95−1.00) | 0.025 |
| Waist Circumference | 1.02 (1.01−1.03) | 0.004 |
| SBP | 1.01 (1.01−1.02) | <0.001 |
| DBP | 0.99 (0.98−0.99) | <0.001 |
| Total Cholesterol | 1.00 (1.00−1.00) | 0.034 |
| Fasting Blood Glucose | 1.00 (1.00−1.00) | <0.001 |
| AST | 1.01 (1.00−1.01) | <0.001 |
| ALT | 0.99 (0.98−0.99) | <0.001 |
| GGT | 1.00 (1.00−1.00) | <0.001 |
| Platelet | 1.00 (1.00−1.00) | 0.014 |
| Total Bilirubin | 1.30 (1.08−1.56) | 0.005 |
| Creatinine | 1.28 (1.13−1.45) | <0.001 |

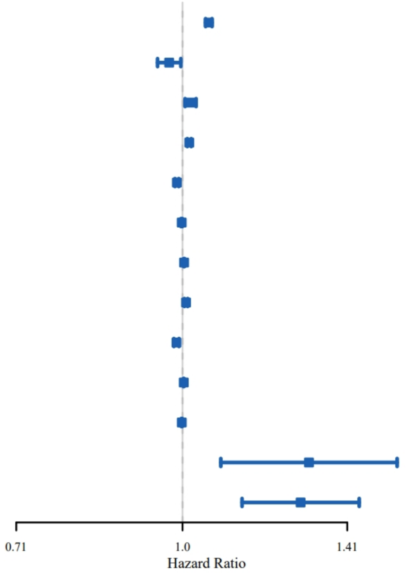


Forest plot displaying adjusted hazard ratios (HR) and corresponding 95% confidence intervals (CI) for independent predictors of all-cause mortality derived from the final multivariate Cox proportional hazards regression model in MASLD patients. Each blue square represents the adjusted HR value of a single variable, and the horizontal line extending from each square denotes its 95% CI. The vertical dashed reference line at HR=1 indicates no significant correlation with mortality risk. Squares and horizontal lines located entirely to the right of the reference line represent risk factors for all-cause death, whereas those fully on the left side indicate protective factors. Abbreviations: HR, hazard ratio; CI, confidence interval; SBP, systolic blood pressure; DBP, diastolic blood pressure; AST, aspartate aminotransferase; ALT, alanine aminotransferase; GGT, gamma-glutamyl transferase.

***Figure S4. Visual validation of the smart data profiling node for metadata aggregation and privacy preservation.***

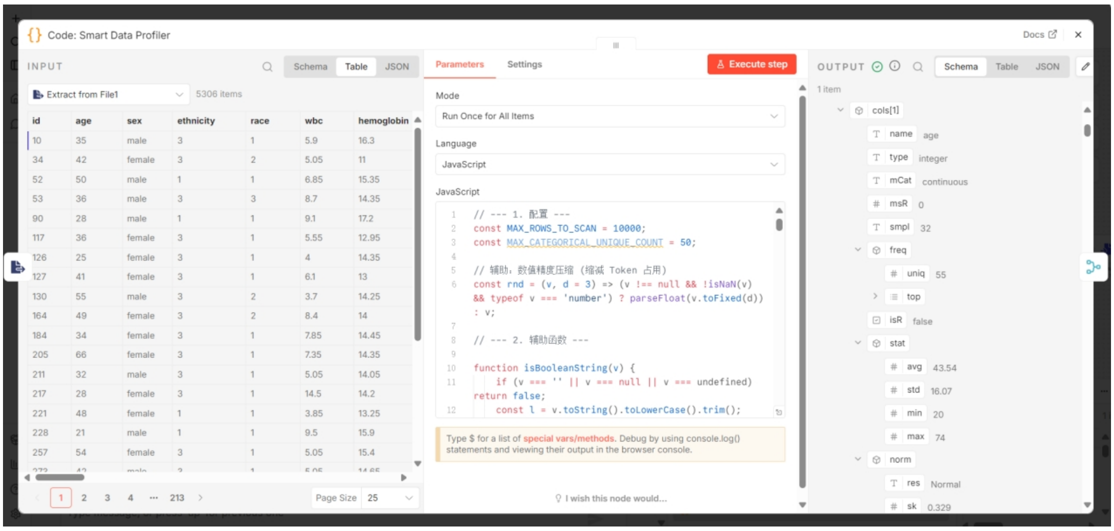


The screenshot captures the operational interface of the Code: Smart Data Profiler node within the local n8n orchestration environment, demonstrating the empirical framework for localized data profiling and privacy isolation.

The interface is structured into three continuous functional panels that validate the secure pipeline before any external LLM interaction:

**INPUT (Left Panel):** Represents the raw clinical research registry residing entirely within the local file system (containing 5,306 item records including patient identifier codes, age, sex, ethnicity, and metabolic/hematologic biomarkers like wbc and hemoglobin). This row-level patient data matrix is restricted to the local compute runtime environment and is completely blocked from outbound network traffic.

**JavaScript Parameters (Center Panel):** Illustrates the localized execution of JavaScript algorithms that process up to 10,000 rows to extract high-level metrics, handle continuous/categorical distribution characteristics, perform decimal precision compression, and evaluate baseline data integrity metrics without relying on cloud-based processing.

**OUTPUT (Right Panel):** Showcases the finalized, aggregated structural metadata object (cols[1] profiling the variable age) generated for downstream reasoning. The extracted properties are strictly restricted to non-sensitive statistical descriptors, including physical/semantic types (integer, continuous), unique value distribution flags (uniq), descriptive central tendencies (avg = 43.54), variance metrics (std = 16.07), boundary ranges (min = 20, max = 74), and normality distribution diagnostics (norm.res = Normal, skewness sk = 0.329).

By demonstrating that the local node extracts and routes only this anonymized, aggregated metadata package to downstream AI agents, the visual workflow provides auditable validation that row-level patient profiles remain natively sandboxed. Abbreviations: LLM, Large Language Model; MAX, Maximum; MIN, Minimum; STD, Standard Deviation.
